# Misclassification of a whole genome sequence reference defined by the Human Microbiome Project: a detrimental carryover effect to microbiome studies

**DOI:** 10.1101/19000489

**Authors:** DJ Darwin R. Bandoy, B Carol Huang, Bart C. Weimer

**Author notes:** corresponding author; (01).

## Abstract

Taxonomic classification is an essential step in the analysis of microbiome data that depends on a reference database of whole genome sequences. Taxonomic classifiers are built on established reference species, such as the Human Microbiome Project database, that is growing rapidly. While constructing a population wide pangenome of the bacterium *Hungatella*, we discovered that the Human Microbiome Project reference species *Hungatella hathewayi* (WAL 18680) was significantly different to other members of this genus. Specifically, the reference lacked the core genome as compared to the other members. Further analysis, using average nucleotide identity (ANI) and 16s rRNA comparisons, indicated that WAL18680 was misclassified as *Hungatella*. The error in classification is being amplified in the taxonomic classifiers and will have a compounding effect as microbiome analyses are done, resulting in inaccurate assignment of community members and will lead to fallacious conclusions and possibly treatment. As automated genome homology assessment expands for microbiome analysis, outbreak detection, and public health reliance on whole genomes increases this issue will likely occur at an increasing rate. These observations highlight the need for developing reference free methods for epidemiological investigation using whole genome sequences and the criticality of accurate reference databases.

## Background

Clostridia are a very diverse group of organisms. The taxonomy is in constant revision in light of new whole genome sequence production and genomic flux^1^. While organism classification can be reassigned, the identified isolates within the same species retain their relatedness. In the analysis of 13,151 microbial genomes, the misclassification (18%) was determined by binning into cliques and singletons with ANI data using the Bron-Kerbosch algorithm, which resulted in the misclassification of 31 out of the 445 type strains^2^. The different causes of the type strain misclassification include poor DNA-DNA hybridization (e.g. high genomic diversity), low DNA-DNA hybridization values, naming without referencing to another type strain, and lack of 16s rRNA data. *Hungatella hathewayi*, or its prior designation *Clostridium hathewayi*, was not included in the previous as there were very few *Hungatella* genomes in the time of that publication. As more metagenomes are published increasing claims of finding new organisms are mounting. To this point, Almeida et al. reported an increase of 1952 uncultured organisms that are not represented in well-studied human populations, where they presented data to support that rare species will be difficult to accurately identify and do not match existing references^3^.

Public repositories of genomic data have experienced tremendous expansion beyond human curatorial capacities, which is an ever increasing issue with the high rate of WGS production^4,5^. Recently, it was estimated that ∼18% of the organisms are misclassified in microbial genome databases^2^. This high rate of error led to investigation of misclassification of specific organisms, including *Aeromonas*^*6*^ *Fusobacterium*^*7*^, and ultimately entire reference databases^2^. These studies found misclassified type strains, which calls into question the foundation of the taxonomy and inferred relatedness when population genomes are being used for epidemiological purposes, especially with rare organisms that are not well represented in the reference database. The work presented here uniquely identified a misclassified reference species and found propagation of incorrectly labelled genomes in several highly cited microbiome studies^8,9,10,11^.

### Observation

Based on this species delineation notion, we discovered that the Human Microbiome Project reference genome for *Hungatella hathewayi* (WAL18680) was misidentified while building a phylogeny of *Hungatella* species using a population of whole genome sequences^12^. Both 16s rRNA and average nucleotide identity (ANI^2^) analysis indicated that WAL18680 was not a member of the *Hungatella* genus based on genome assessment (Table 1). Population genome comparison analysis was instrumental in discovering that WAL18680 was misclassified and the impact for genomic epidemiology purposes would be important.

**Table 1.** Average nucleotide identity (ANI) of *Hungatella* isolates using the WGS. The reference WGS from WAL18680 (isolated in Canada in 2011) was classified as a different genus using the ANI criteria as compared to the other isolates examined. Strains 2789STDY5834916 (isolated in the UK in 2015) and BCW8888 (isolated in Mexico in 2015) would be considered new and novel species using ANI.

|  | *H. hathewayi*<br>BCW8888 | *H. effluvii*<br>DSM24995 | *H. hathewayi*<br>WAL18680 | *H. hathewayi*<br>VE202-11 | *H. hathewayi*<br>2789STDY5834916 | *H. hathewayi*<br>2789STDY5608850 | *H. hathewayi*<br>12489931 |
| --- | --- | --- | --- | --- | --- | --- | --- |
| *H. hathewayi*<br>BCW8888 | 100 | 94.3 | 70.9 | 96.7 | 85.4 | 98.1 | 96.8 |
| *H. effluvii*<br>DSM24995 | 94.4 | 100 | 70.5 | 94.5 | 85.5 | 94.4 | 94.9 |
| *H. hathewayi*<br>WAL18680 | 71.2 | 70.8 | 100 | 71.0 | 72.0 | 71.1 | 71.1 |
| *H. hathewayi*<br>VE202-11 | 96.6 | 94.5 | 70.7 | 100 | 85.2 | 96.5 | 98.73 |
| *H. hathewayi*<br>2789STDY5834916 | 85.6 | 85.6 | 72.0 | 85.4 | 100 | 85.7 | 85.8 |
| *H. hathewayi*<br>2789STDY5608850 | 98.1 | 94.3 | 70.7 | 96.4 | 85.4 | 100 | 96.5 |
| *H. hathewayi*<br>12489931 | 96.8 | 94.9 | 70.8 | 98.8 | 85.7 | 96.7 | 100 |

The misclassified *H. hathewayi* WAL18680 has been used to generate phylogenomic analysis, reference WGS for metagenome analysis, and web server identification platforms utilizing the metagenomic classifiers^10,13,14^. Epidemiologically, association with clinical disease will be discordant with genomic data and result in inaccurate conclusions on the microbiome ecology or therapies based on the microbiome membership to mitigate disease leading to the wrong causal relationship to be concluded^9^. As more microbiome studies are linking rare microbes to biological outcomes, a need exists to quickly identify inaccurate assignment when only a few WGS of individual organisms are available for use as a reference. This creates an issue with low sampling of the genome space for rare organisms and may result in mis-naming based on a small set of phenotypic assays that do not represent the genome content or flux^15^.

*H. hathewayi* was first described as an isolate was from human feces^16^ and was subsequently reported in a patient with acute cholecystitis, hepatic abscess, and bacteremia^17,18^. It was also later reported in a case of appendicitis^19^. *H. hathewayi* is (WAL18680) one of the designated reference strains in Human Microbiome Project and is used extensively for binning and classification of microbiome related studies, which confounds analysis of the genus *Hungatella*. This organism can be isolated from the microbiome depending on the enrichment conditions^9^. Having a reference species misclassified is detrimental to microbiome research and in epidemiological investigations. To solve this issue, we developed a heurist to minimizing misclassification for rare reference species as a result of cross-validation of the genomic information for name assignment.

The standard procedure of the 100K Pathogen Genome Sequencing Project^4,5,20-22^ determines the identity of bacterial pathogen isolates in clinical samples using WGS and the genome distance (ANI^23,24^) before proceeding with additional comparisons. This analysis was done with a group of isolates from suspected *Clostridioides difficile* infection cases. We identified a species of *H. hathewayi* using genome distance using the entire genome sequence that was implemented for high dimensional comparison using MASH^25^ (with the maximum sketch size). This was coupled to comparison of all of the available WGS to represent the entire genome diversity to build a whole genome phylogeny^12^ to determine the naming accuracy of the clinical isolates. Unexpectedly, one particular sequence was well beyond the species ANI threshold for *C. difficile*. We found that based on ANI, is a putative new species of *Hungatella* (strain 2789STDY5834916). Weis et al.^26,27^ used this method with *Campylobacter* species to demonstrate that genome distance accurately estimates host-specific genotypes, zoonotic genotypes, and disease within livestock disease with validated reference genomes. While ANI was the first estimate to raise questions for the accurate identification of this organism, we proceeded with a cross-validation strategy to verify the potential misclassification of the reference species.

We advanced with the initial mis-identification by determining the pangenome analysis with the hypothesis that outbreak isolates would cluster together based on the isolate origin (i.e. an individual or location)^*12*^ as well as contain the same core genome. We found that WAL18680 did not contain any of the core genome relative to all of the other *Hungatella* genomes (Figure 1). Together, these genomic metrics prove that this reference genome was misclassified, which has extensive implications as reference sequences are commonly used for genomic identity for outbreak investigations. Additionally, metagenome studies require reference genome databases to identify bacterial community members. This result indicates that if the epidemiological workflow did not include specific whole genome alignment, inaccurate conclusions and misleading deductions will be made – as was observed by Kaufman et al.^15^ – where they found that genome diversity is unexpectedly large and expands based on a power law with each new WGS that is added to the database. Combining the fact that this is a reference genome from a rare organism from a very diverse group, that the genome evolution rate is a power law, and that this is a reference genome from the Human Genome Project the implications for the mis-identification have far reaching implications.

**Fig 1.**
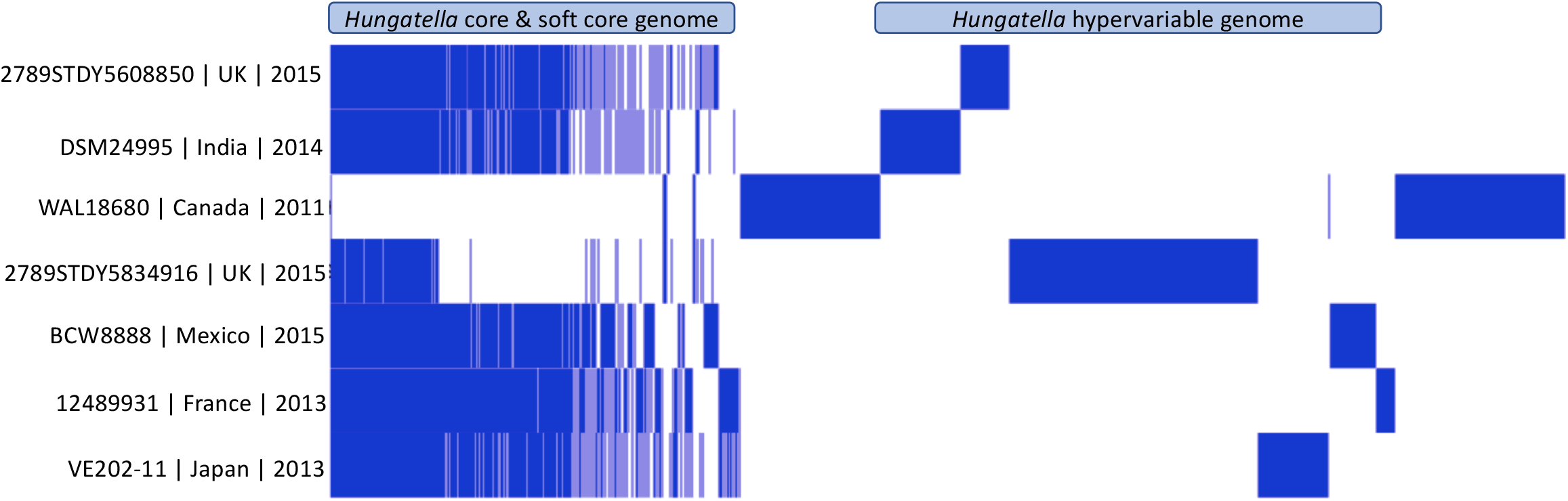
Pangenome of *Hungatella*. WAL18680 was originally identified as ***Clostridium hathewayi***. After a recent taxonomic reclassification it was renamed as *Hungatella hathewayi*. **(WAL 18680) does not have the core genome of other *Hungatella* species (***hathewayi* ***or*** *efluvii)* and possess very few core genes common to the other *Hungatella* species. The bulk of its genome is not found in other *Hungatella* species, indicating it belongs to another genus. Strain 2789STDY5834916 is a novel *Hungatella* species.

**Fig 2.**
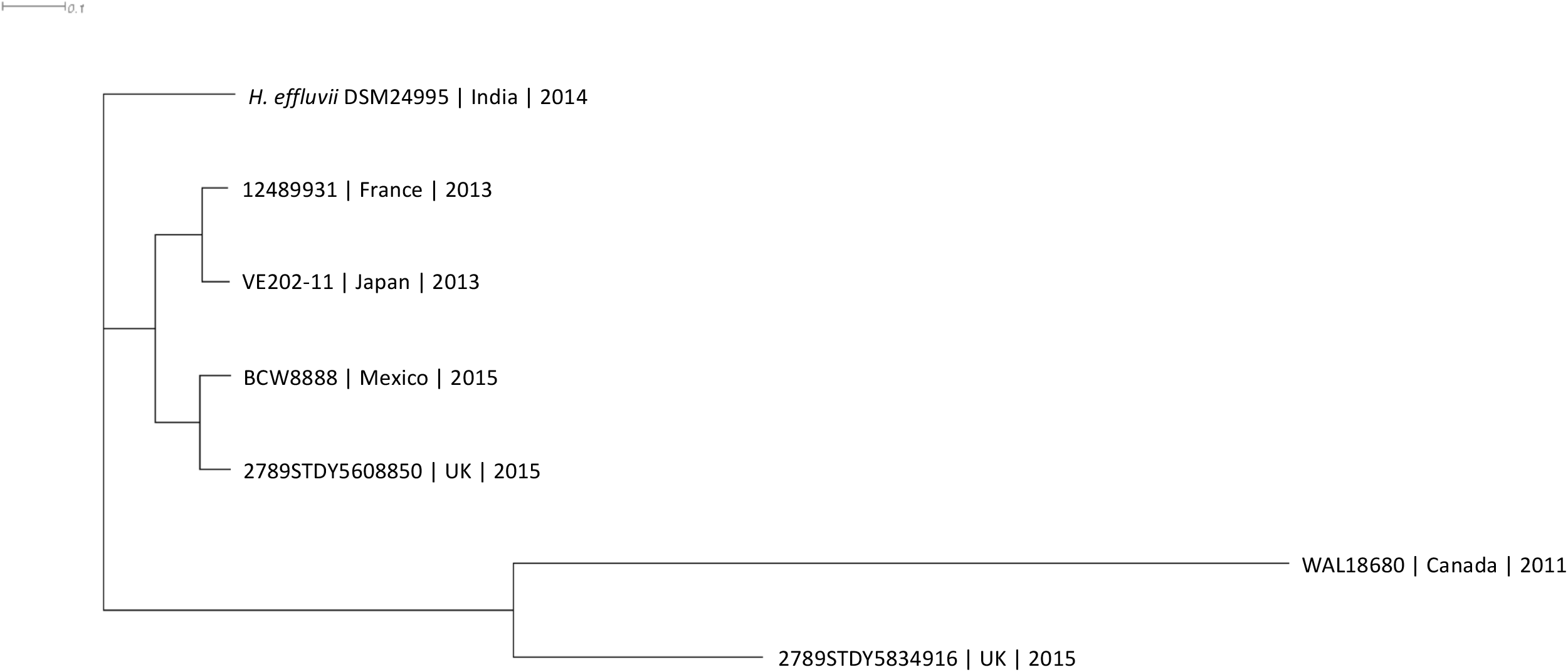
Phylogenomic of all *Hungatella* relatedness estimated using genome distance.

Conflicts of taxonomic classification based on traditional methods, such as phenotypic assays, metabolism, with genomic based parameters will likely increase as more genomes are produced and use of the entire genetic potential (i.e. the entire genome). The need for heuristical indicators of misclassification are needed as is the need to expand WGS that adequately represent bacterial diversity among and within taxonomy to represent the genetic diversity of any single organism.

### Genome sequence availability

The WGS for each genome is via the NCBI with Biosample numbers of SAMD00008809, SAMN02463855, SAMN02596771, SAMEA3545258, SAMEA3545379, SAMN09074768. The WGS sequence for BCW8888 is available via the 100K Project BioProject at the NCBI (PRJNA186441) as Biosample SAMN12055167.

## Data Availability

The whole genome sequences are available now via the SRA for all bu, except BCW8888. It will be publically available within 90 days.

